# The respiratory sound features of COVID-19 patients fill gaps between clinical data and screening methods

**DOI:** 10.1101/2020.04.07.20051060

**Authors:** Yinghui Huang, Sijun Meng, Yi Zhang, Shuisheng Wu, Yu Zhang, Yawei Zhang, Yixiang Ye, Qifeng Wei, Niangui Zhao, Jianping Jiang, Xiaoying Ji, Chunxia Zhou, Chao Zheng, Wen Zhang, Lizhong Xie, Yongchao Hu, Jianquan He, Jian Chen, Wangyue Wang, Liming Cao, Wen Xu, Yunhong Lei, Zhenghua Jiang, Weiping Hu, Wenjuan Qin, Wanyu Wang, Yulong He, Hang Xiao, Xiaofang Zheng, Yiqun Hu, Wensheng Pan, Changhua Zhang, Jianfeng Cai

## Abstract

**Background:** The 2019 novel coronavirus (COVID-19) has continuous outbreaks around the world. Lung is the main organ that be involved. There is a lack of clinical data on the respiratory sounds of COVID-19 infected pneumonia, which includes invaluable information concerning physiology and pathology. The medical resources are insufficient, which are now mainly supplied for the severe patients. The development of a convenient and effective screening method for mild or asymptomatic suspicious patients is highly demanded.

**Methods:** This is a retrospective case series study. 10 patients with positive results of nucleic acid were enrolled in this study. Lung auscultation was performed by the same physician on admission using a hand-held portable electronic stethoscope delivered in real time via Bluetooth. The recorded audio was exported, and was analyzed by six physicians. Each physician individually described the abnormal breathing sounds that he heard. The results were analyzed in combination with clinical data. Signal analysis was used to quantitatively describe the most common abnormal respiratory sounds.

**Results:** All patients were found abnormal breath sounds at least by 3 physicians, and one patient by all physicians. Cackles, asymmetrical vocal resonance and indistinguishable murmurs are the most common abnormal breath sounds. One asymptomatic patient was found vocal resonance, and the result was correspondence with radiographic computed tomography. Signal analysis verified the credibility of the above abnormal breath sounds.

**Conclusions:** This study describes respiratory sounds of patients with COVID-19, which fills up for the lack of clinical data and provides a simple screening method for suspected patients.

## Introduction

The 2019 novel coronavirus (COVID-19) broke out in Wuhan in January 2020, which has now spread all over the world^1,2^. Respiratory droplets and close contacts are the main transmission routes of COVID-19^3^. Cough was the first symptom in 67.8% of COVID-19 infected patients^4^.

Currently, the identification of suspected patients is mainly based upon epidemiological data and clinical symptoms. With the occurrence of pandemic, more and more patients without a clear epidemiological history or typical clinical symptoms have been found. A more convenient and more reliable method of identifying suspicious patients is urgently needed. The main laboratory examination methods are nucleic acid testing and chest imaging^5^. But the former is a diagnostic test, which may be priority to the suspected patients after primary screening. The latter is expensive and difficult to obtain in most countries. At the same time, both of them need the interval time waiting for results, which extends the time to take measures such as isolation.

Pulmonary auscultation^6,7^ is an important means to diagnose and treat pulmonary diseases in the routine medical work, which can provide important physiological and/or pathological information. However, various causes limited its application when faced with infectious diseases. Electronic stethoscope can transmit auscultation data through Bluetooth in real-time^8,9^. It can break the status quo of unable-auscultation caused by medical protective clothing without increasing the risk of medical infection. This study intends to leverage a remote electronic stethoscope to collect the lung auscultation characteristics of COVID-19 patients, which makes up the lack of existing COVID-19 clinical data. It also proposes to use remote equipment in screening and monitoring patients with COVID-19, which is a safe and effective way to break the regional restrictions.

## Methods

### Data source

The case series with COVID-19 were from the Affiliated Nanping First Hospital of Fujian Medical University (NPFH), located in Nanping, Fujian Province. NPFH was the designated hospital for admission of COVID-19 infection patients in Nanping city, and set up the expert group for medical treatment of COVID-19 (Conv-MG). Suspicious patients found by other medical units in Nanping city after preliminary screening should be transferred to NPFH for treatment. All patients with NCIP enrolled in this study were diagnosed according to World Health Organization interim guidance. From January 29^th^ to February 14^th^, the ConV-TG of NPFH used electronic stethoscopes to auscultate the lungs of newly admitted patients with COVID-19 infection. On March 5, after all the patients were discharged from the hospital, the clinical data and laboratory examination results of the above patients were retrospectively analyzed, which included the pulmonary signs.

The study was approved by the ethics committee of NPFH (202002050). Oral permission was obtained from patients before lung auscultation. All patients were informed prior to physical examination. With the patient’s oral consent, we gave them lung auscultation.

### Electronic stethoscope, auscultation and recording method

The stethoscope utilizes Doctorgram stethoscope DES-I (GV concepts, USA), which is a Hand-held portable Bluetooth electronic stethoscope. The stethoscope head is membrane-shaped and bell-shaped, and the frequency range of receiving signals is 20Hz-2KHz, which is amplified for 24 times. There is no separate filtering of cardiac and respiratory sounds. The signal is transmitted to the software terminal of the mobile phone or computer in real time through Bluetooth. The number of terminals is unlimited, and multiple doctors can be connected at the same time. The sound can be stored by the lossless compression technology of FLAC, which can continue record for 30 s.

In this study, respiratory sound data were acquired on the day of patients were admitted to the NPFH. All records were obtained with the subjects in a seated position. The order of auscultation starts from the tip of the lung, from top to bottom. The anterior chest, lateral chest and back are symmetrically examined from left to right. Tracheal breath sounds, bronchial breath sounds, bronchoalveolar breath sounds and alveolar breath sounds were examined and collected respectively (Figure S1.). Lastly, speech resonance is detected and left-to-right contrast is made. All breath sounds of each patient were collected by the same physician in the isolation ward, to ensure the stability of audios collected from different patients. The Conv-MG conducted multidisciplinary treatment (MDT) by video equipment outside of the isolation ward.

The exported audio is preliminarily reviewed and checked by the doctor who collected the breath sounds. To conduct auscultation inspection on the recording quality, sort and label the audio by the corresponding auscultation position as well as the type of breath sound. Another data analyst used audio editing software (Cool Edit Pro v. 2.0; Syntrillium Software Corporation, USA) to initially edit the audio according to the previous label, and remove the audio with low quality. Exclusion criteria include: audio with high background noise, short recording time (at least 2 complete breathing sounds), and low breathing sound signal due to poor contacts or conduction.

### Methods of auscultation recording analysis

The audio analysis was conducted by six respiratory specialists having 5 to 23 years of experience. The six doctors are from five hospitals, including NPFH, the Seventh Affiliated Hospital of Sun Yat-sen University, Pucheng County Hospital of Traditional Chinese Medicine, First Affiliated Hospital of XiaMen University, and the Second Afficiated Hospital of Xiamen Medical College. There was no communication between these physicians before and after auscultation. The content of auscultation assessment includes breathing rate, intensity, and whether there were abnormal or additional breath sounds. Each audio was played over and over again until the physician made a decision. All the results of each physician were recorded in sheet, then summarized and normalized by two graduate students who were blinded to patient information. The terminology proposed by the ad hoc committee of the International Lung Sounds Association was used to agree on the respiratory sounds described by six physicians.

In addition to the result judgments of respiratory doctors, the above audio is also graphically displayed and quantitatively described by means of signal analysis using Matlab R2019, v9.6.0 (MathWorks, USA). The relationship between signal characteristics and time is intuitively demonstrated by drawing waveform and time-frequency diagram. By observing the waveform of breath sound, the duration of inhalation and breath as well as the intensity of sound can be preliminarily understood. The main frequency range of normal and abnormal breath sounds can be visualized by detecting the time-frequency chart. The background noise generated by the movement between the patient and the stethoscope is a high energy artifact on the time-frequency graph, which can be easily recognized in the image. The feature of normal respiratory sounds and pneumonic respiratory sounds refer to APSP project^10^. The images were excluded from the analysis. Then the corresponding spectrum diagram was drawn by fast Fourier transform (FFT), which can analyze the signal strength of a particular frequency.

### RT-PCR testing and chest radiographic computed tomography (CT) examination

Throat swabs were taken for each patient, and the samples were delivered to the designated detection institution immediately after collection. The detection kit was COVID-19 RT-PCR Kit (Shanghai ZJ Bio-tech, China), which target E gene, RdRp gene, and N gene. All patients underwent chest CT examination after admission through a CT scanner (General Electric Company, USA). The scanning range was from the level of the thoracic inlet to the level of the diaphragm. Lung window and mediastinal window were collected, and the CT image characteristics of patients were analyzed by two experienced imaging physicians on the workstation of picture archiving and communication systems.

### Statistical analysis

Categorical variables were described as frequency rates and percentages, and continuous variables were described using median, and distribution range. These statistical analyses were performed using SPSS version 13.0 (SPSS Inc, USA).

## Results

### Characteristics of the patients

A total of 10 patients were included in this study, ranging in age from 30 to 82 years old, mostly male (70%), all of whom had an epidemiological history of contact with suspected or confirmed patients. The number of patients was limited since current situation was serious and it was safe to detect the results through patients in one ward. The most common initial symptoms were similar to recent studies, which included respiratory symptoms (6/10) or fever (1/10) with highest temperature reaching 39.2°C. One patient was asymptomatic and had only transient fever during treatment, with a maximum temperature of 37.5°C. Two other patients had the first symptoms of digestive system, one of which was diarrhea and the other was abdominal discomfort. However, both patients developed cough and fever successively before admission. Before admission to NPFH, all patients were isolated and observed in regional primary medical institutions. All patients had at least one nucleic acid test, and 7 patients had a positive test at the first time, the other 3 patients had a negative test repeated the test after 24 hours. The median incubation period from primary symptoms to viral test positive was 7 days (interquartile range 3-15 days). Once the nucleic acid test is found positive, the patient was immediately transferred to COV-MG of NPFH for intensive treatment. Chest X-ray or CT were detected before and after admission for each patient. Local or bilateral ground glass shadows, infiltrating shadows or stromal changes were found in all patients’ CT, of which 80% patients accumulated bilateral lungs. Based on further examination which were done after admission, three patients were diagnosed with severe type and seven patients were diagnosed as common type, all of whom did not develop to comorbid conditions.

### Respiratory sound features of COVID-19 infected patients

Common abnormal breath sounds include bronchial breathing (common in pneumonia or fibrosis), stridor (indicating upper-airway obstruction), wheeze (typical signs of asthma), rhonchus (common with airway narrowing caused by bronchitis, chronic obstructive pulmonary disease), cackles (including fine cackles and coarse, can be earliest sign of disease, eg. interstitial lung fibrosis, congestive heart failure, pneumonia, asbestosis, etc.), pleural friction rub (associated with pleural related disease), squawk (associated with conditions affecting distal airways)^6^. After the preliminary judgment of breath sounds collected from ten patients, two patients with poor recording quality were excluded according to exclusion criterion. None of the patients had a history of chronic respiratory disease, which may interference with auscultation results. Among the results of breath sounds auscultation from six doctors, the types of abnormal breath sounds described by each doctor had some differences. These were related to factors such as the auscultation habits of each doctor.

Among them, asymmetric voice tremor and rales were the abnormal breath sounds that were easy to detect. For each patient, only one or more abnormal breath sounds were described in each doctor’s auscultation results, in which the patient was recorded as abnormal breath sounds and the diagnosis was positive. Based on the above method, 1 of the 8 patients was diagnosed with abnormal breath sounds by 6 physicians at the same time. 2 patients were diagnosed by 5 physicians at the same time, 4 patients were diagnosed by 4 physicians at the same time, and only 1 patient was found to have abnormal breath sounds by 3 physicians (Table 2).

**Table 1.** Basic information of patients.

| **Patients (n=10)** | **All** | **First episode of**<br>**respiratory**<br>**symptoms**<br>**(n=7)** | **First episode of**<br>**digestive**<br>**symptoms**<br>**(n=2)** | **Asymptomatic**<br>**(n=1)** |
| --- | --- | --- | --- | --- |
| **Age, years** |  |  |  |  |
| Range | 30-82 | 30-53 | 58-82 | 38 |
| **Sex** |  |  |  |  |
| Female | 7(70%) | 5(71%) | 1(50%) | 1(100%) |
| Male | 3(30%) | 2(29%) | 1(50%) | 0(0) |
| **Exposure to source of**<br>**transmission within past 14**<br>**days** | 10(100%) | 7(100%) | 2(100%) | 1(100%) |
| **Signs and symptoms at**<br>**admission** |  |  |  |  |
| Fever | 9(90%) | 7(100%) | 2(100%) | 0(0) |
| Cough | 9(90%) | 6(86%) | 2(100%) | 0(0) |
| Expectoration | 5(71%) | 5(71%) | 0(0) | 0(0) |
| Shortness of breath | 3(30%) | 2(29%) | 1(50%) | 0(0) |
| Muscle ache | 1(10%) | 1(14%) | 0(0) | 0(0) |
| Fatigue | 1(10%) | 1(14%) | 0(0) | 0(0) |
| Headache | 0(0) | 0(0) | 0(0) | 0(0) |
| Sore throat | 0(0) | 0(0) | 0(0) | 0(0) |
| Rhinorrhoea | 2(20%) | 1(14%) | 1(50%) | 0(0) |
| Chest pain | 0(0) | 0(0) | 0(0) | 0(0) |
| Diarrhoea | 1(10%) | 1(14%) | 0(0) | 0(0) |
| Nausea and vomiting | 1(10%) | 0(0) | 1(50%) | 0(0) |
| More than one sign | 8(90%) | 6(86%) | 2(100%) | 0(0) |
| **Median incubation time**<br>**between admission symptom**<br>**to nucleic acid test positive**<br>**(days)** | 7(3-15) | 7(3-15) | 8.5(6,11) | 0(0) |
| **Nucleic acid test** |  |  |  |  |
| Positive for the first time | 7(67%) | 5(70%) | 1(50%) | 1(100%) |
| Positive for the second time | 3(33%) | 2(30%) | 1(50%) | 0(0) |
| **Chest x-ray and CT findings** |  |  |  |  |
| Groud-glass opacity | 7(70%) | 5(71%) | 1(50%) | 1(100%) |
| Bilateral patchy shadowing | 2(20%) | 2(29%) | 0(0) | 0(0) |
| Unilateral patchy shadowing | 8(80%) | 5(71%) | 2(100%) | 1(100%) |
| Interstitial abnormalities | 8(80%) | 5(71%) | 2(100%) | 1(100%) |
| **Chronic medical illness** |  |  |  |  |
| Cardiovascular disease | 2(20%) | 1(14%) | 1(50%) | 0(0) |
| Digestive system disease | 1(10%) | 1(14%) | 0(0) | 0(0) |

**Table 2.** Diagnosis results of lung sound.

|  | Fever | Respiratory symptom | X-ray or CT results | Auscultation results by respiratory physicians |  |  |  |  |  |
| --- | --- | --- | --- | --- | --- | --- | --- | --- | --- |
|  |  |  |  | Physician 1 | Physician 2 | Physician 3 | Physician 4 | Physician 5 | Physician 6 |
| Patient1 | No | No | Bilateral patchy pneumonia | Normal | #Cackles, *Avr | *Avr-m | Normal | *Vbs-m, *Avr-m | Avr |
| Patient2 | No | Yes | Bilateral patchy pneumonia | Normal | *R-Vbs-m | *R-Vbs | Normal | Vbs-m, Avr-m | Normal |
| Patient3 | Yes | Yes | Bilateral patchy pneumonia | Avr | Cackles, Avr-m | Normal | Cackles, Avr-m | Vbs-m, Avr-m | Cackles, Avr-m |
| Patient4 | Yes | Yes | Bilateral patchy pneumonia | / | / | / | / | / | / |
| Patient5 | Yes | Yes | Bilateral patchy pneumonia | Normal | R-Vbs-m, Avr-m | *Vbs | Normal | Vbs-m, Avr-m | Vbs-m, Avr-m |
| Patient6 | Yes | Yes | Bilateral patchy pneumonia | Cackles | Cackles, Avr-m | Vbs, Avr-m | Avr | Cackles, Avr-m | Vbs-m, Avr-m |
| Patient7 | No | Yes | Bilateral patchy pneumonia | / | / | / | / | / | / |
| Patient8 | Yes | Yes | Right pneumonia | Normal | Cackles, *Vr-m | Normal | Cackles | Vbs-m, Avr-m | Avr |
| Patient9 | Yes | No | Right pneumonia | Cackles | Cackles, Avr | Avr-m | Normal | Avr-m | Avr-m |
| Patient10 | Yes | Yes | Bilateral patchy pneumonia | Normal | Cackles, Avr | Avr | Normal | Avr-m | Avr-m |

Vr: vocal resonance; Avr: Asymmetric vocal resonance; Vbs: Vesicular breath sounds; R-Vbs: Reduced vesicular breath sounds; Vr-m: vocal resonance with murmurs; Avr-m: Asymmetric vocal resonance with murmurs

The diagnosis results of respiratory sounds in the asymptomatic patients included cackles, asymmetric vocal resonance, and abnormal vesicular breath sounds with murmurs. Asymmetric vocal resonance with the highest diagnostic consistency amongst 6 physicians. We use signal analysis to quantitatively describe this abnormal breath sound. After extracting the audio of bilateral vocal resonance in the anterior chest (Figure 1.A-B), we drew the waveform and spectrum (Figure 1.C). It can be seen that the frequency distribution of breath sounds on both sides is similar, but the left breath sound intensity significantly increases, which is consistent with the CT results (See Figure 1. for more details).

**Figure 1.**
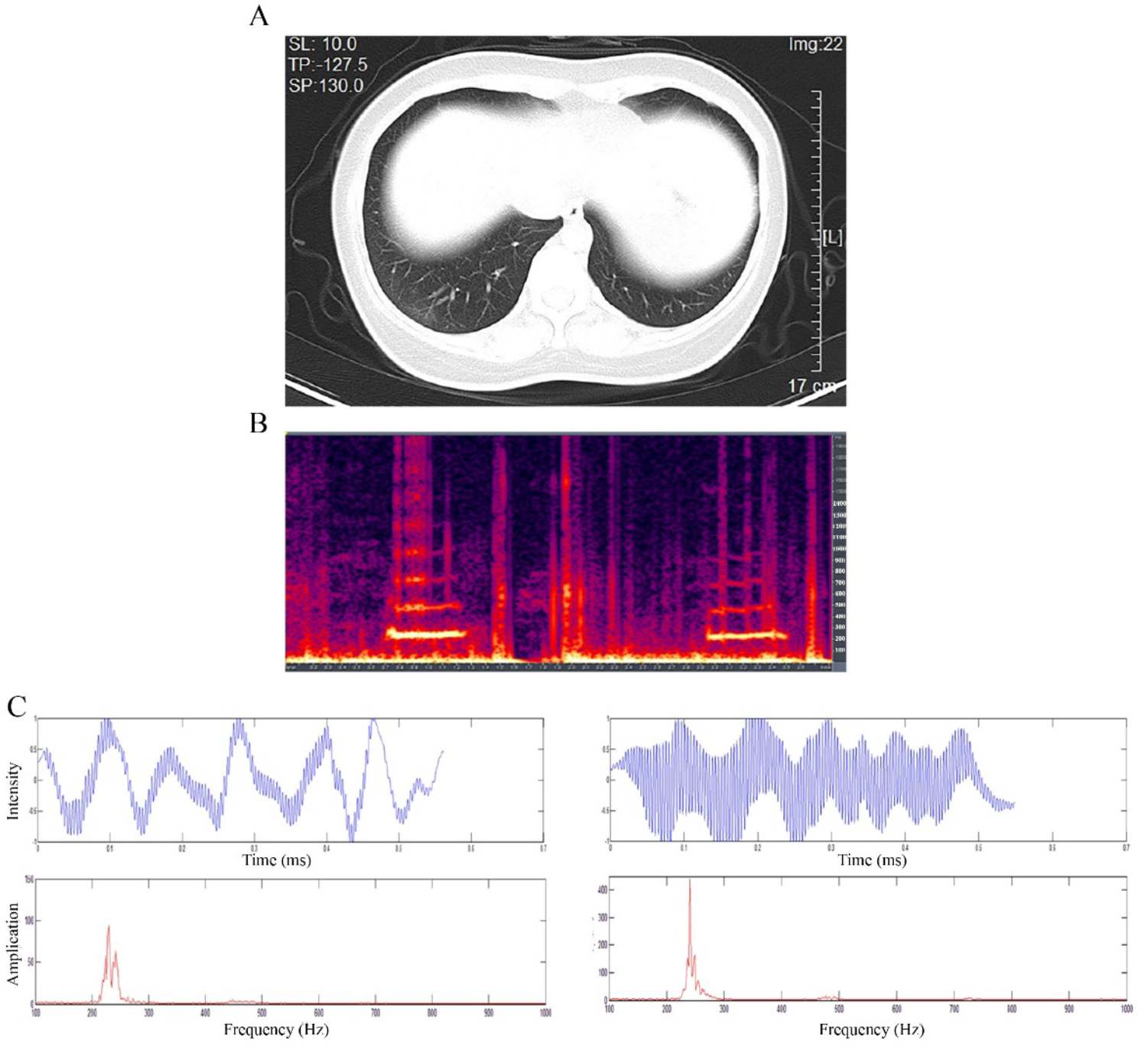
CT image and vocal resonance signal feature of asymptomatic patients. A. The image shows the CT scan of the 9th intercostal cross section of the asymptomatic patient. The increased density of ground glass in the subpleural area of double lobes was seen, which was considered to be infectious lesions. B. The time-frequency diagram of vocal resonance (collected in the midline of the clavicle meets the 4th intercostal junction), the yellow and red horizontal stripes is the voice print stripe. The stronger the sound, the brighter the color, and vice versa. The yellow and red horizontal stripes in left side corresponds to the right pulmonary voice conduction in figure A, and the yellow and red horizontal stripes in right side corresponds to the left pulmonary voice conduction in figure A. C. The waveform and spectrum of bilateral speech resonance are extracted separately. The right side corresponds to the left chest, and the left side corresponds to the right chest. The blue voiceprint, the vertical axis is sound intensity, and the horizontal axis is time, showing the relationship between sound intensity and time. The graph of the red line, frequency on the horizontal axis and intensity on the vertical axis, shows the distribution frequency of a particular voiceprint. Both sides of the voice resonance frequency is similar, the intensity is different, the left chest is stronger.

Except for asymmetric vocal resonance, cackles were another common abnormal breath sound according to the auscultation results (Table 2). We quantified the signature of one signal of cackles, which was diagnosed by three physicians as a group, to validate the physicians’ auscultation results. The feature of waveform and time-frequency graph (Figure 2.B-C) are consistent with previous description concerning cackles features: duration <100ms, and the main frequency range is about 200-600Hz.

**Figure 2.**
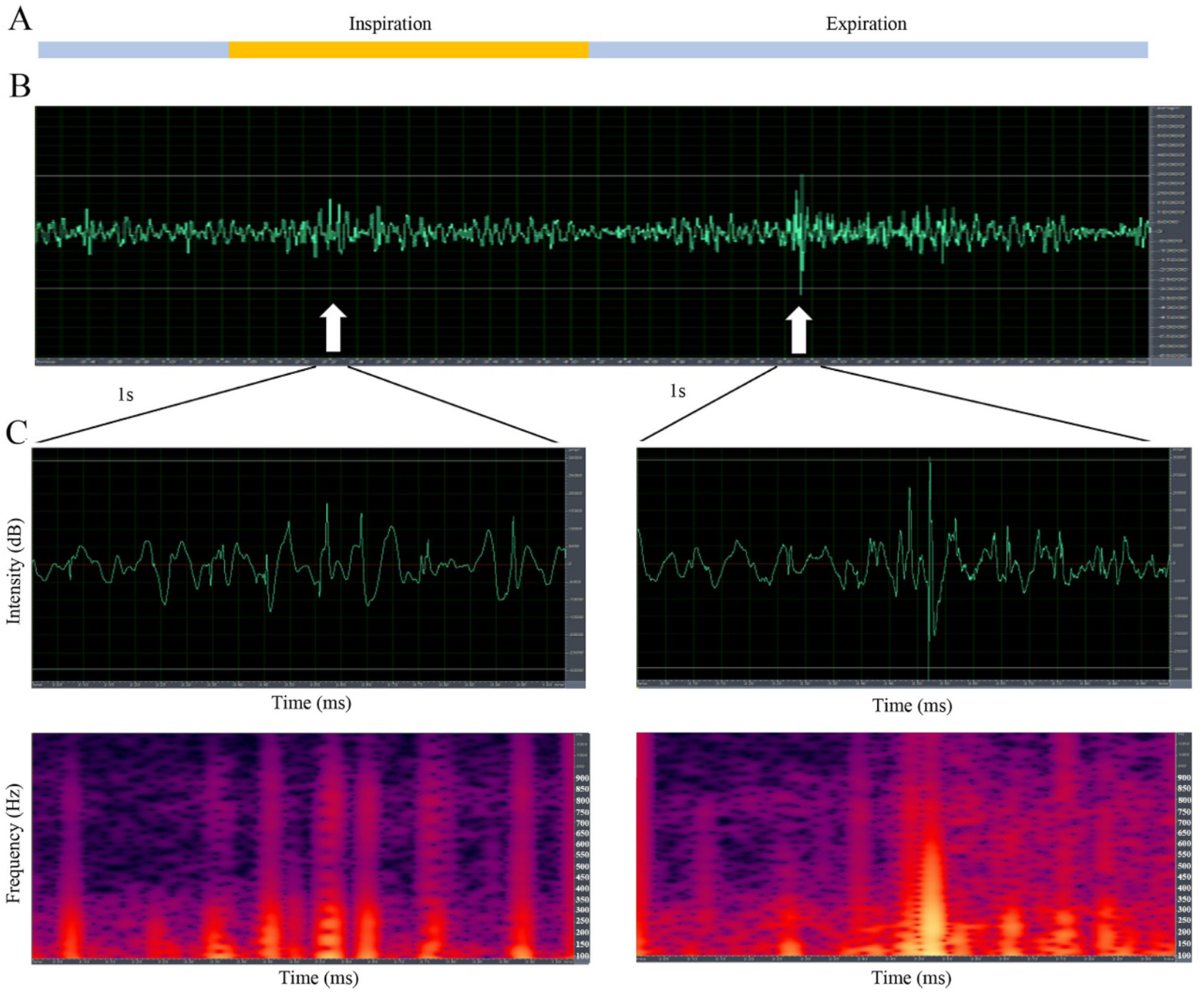
Feature analysis of cackles. A. Ratio of inhalation to exhalation time. Yellow is for breathing in and blue is for breathing out. B. The vesicular breath sounds of the above respiratory cycle. The two white arrows indicate the location of the cackles. C. Extract and amplify the breath sound including cackles to make waveform and time - frequency diagrams respectively. The waveform shows a sharp peak in high intensity range, and the interval time are about 50ms. The main frequencies are between 150 and 600.(Cackles: Crackles are short, explosive, nonmusical sounds heard on inspiration and sometimes during expiration. The feature is duration<100ms, frequency range is about 350Hz (Fine cackles), or 650Hz (Coarse cackles)^6^, other studies have reported the rang is 100-1000 Hz^8^. The likey mechanism of crackles is the sudden inspiratory opening of small airways held closed by surface forces during the previous expiration, which can be correlated to COPD, pneumonia, fibrosis or bronchiectasis^18^.)

## Discussion

The respiratory sounds of 10 patients with COVID-19 infection were recorded and analyzed by electronic stethoscope in this research. Except for the two patients with poor audio quality, all other patients were found to have abnormal breath sounds, including cackles, unable to classified murmurs, abnormal vesicular breath sounds, enhanced or weakened voice resonance. Recent pathological studies indicated that the lungs of patients with COVID-19 presented different degrees of consolidation^11,12^. The imaging examination results suggested that multiple pulmonary plaques, ground glass shadows and infiltrating shadows were the main manifestations of COVID-19 patients, and in severe cases, lung consolidation occurred^4,13^. The auscultation results of patients with COVID-19 infected pneumonia was concordant with the pathological and imaging results, which indicated that the pulmonary signs of patients infected with COVID-19 were not specific compared with other pulmonary inflammation.

However, auscultation can be used as a simple monitoring and screening method to effectively obtain the physiological and pathological information of patients, and no other clinical procedure matches auscultation, providing clinical information about the respiratory system quickly, easily, and almost universally available methods. At present, the focus of pulmonary examination is on X-ray or CT. However, the development of acoustic devices may provide an indirect-contact, credible, repeatable way to monitor the patient’s pulmonary lesions. With the above equipment, the physician wearing protective clothing can collect the lung sound of patient infected COVID-19 without increasing the potential infection rate of his own, and analyze the stored lung sound just as any other clinical information is measured and stored. Existing electronic stethoscopes can already be combined with remote diagnosis and treatment, and automatically detect abnormal respiratory sounds through machine learning^9^. Using this technology to screen patients infected with COVID-19 can improve the recognition rate of infected patient without bringing additional pressure on the use of medical resources.

Another remarkable output is the positive auscultation results of two atypical patients and one asymptomatic patient, which are consistent with the chest CT findings of these patients. Recently, mild or asymptomatic patients with COVID-19 infections attract more attentions^14^. A report based on the passengers of Diamond Princess cruise ship indicate that 4 of the 13 evacuees were asymptomatic patients with positive nucleic acid^15^. Another study found that 56 percent infected children had mild or asymptomatic symptoms^16^. However, these patients showed no reduction in the ability to transmit the virus comparing to typical patients^17^. In the current global pandemic situation, medical resources are scarce in all countries. At present, the initial screening of suspected patients is mainly based on the exposure history of confirmed patients, and it is impossible to conduct large-scale nucleic acid testing or chest imaging screening. However, with the exponential increase of cases, the disease surveillance capacity is limited, and the epidemiological history of many populations is not clear. The accumulation of asymptomatic patients makes it even difficult to control disease. Based on the results of our case-series research, the application of auscultation with electronic stethoscope can make up the shortcomings of current screening methods so as to effectively improve the recognition rate of suspicious patients.

At the same time, there are limitations in this study. Since the results of auscultation are mostly based on personal experience, we did not perform statistical analysis such as the sensitivity, specificity and other indicators of diagnostic test due to lacking effective gold standard. In addition, this is a small observed research, and only the lung breath sounds of the patients are collected at the time of admission. No follow-up research was conducted.

This investigation complements the existing clinical data of COVID-19, which provides a convenient screening method. It is hoped that public health workers around the world will be able to use electronic stethoscope to increase the detection rate of mild and asymptomatic patients, reduce their own infection risk, and allow remote treatment to help fight the epidemic.

## Data Availability

The case series with COVID-19 were from the Affiliated Nanping First Hospital of Fujian Medical University (NPFH), located in Nanping, Fujian Province. NPFH was the designated hospital for admission of COVID-19 infection patients in Nanping city, and set up the expert group for medical treatment of COVID-19 (Conv-MG). Suspicious patients found by other medical units in Nanping city after preliminary screening should be transferred to NPFH for treatment. All patients with NCIP enrolled in this study were diagnosed according to World Health Organization interim guidance. From January 29th to February 14th, the ConV-TG of NPFH used electronic stethoscopes to auscultate the lungs of newly admitted patients with COVID-19 infection. On March 5, after all the patients were discharged from the hospital, the clinical data and laboratory examination results of the above patients were retrospectively analyzed, which included the pulmonary signs.

## Acknowledgement

Thanks to Hesong Qiu for managing and optimizing the research program, to Dr. Teng Hong Dar and Mr. Ching Kay Chow for providing hardware technical support, to Dr. Yonghe Zhou and Mr Kunping Huang for policy coordination assistance. Thank you to every brave warrior fighting this epidemic around the world for your courage.

## Contributors

Yinghui Huang was the physician for auscultation examination. Sijun Meng were responsible for project optimization, data analysis and article written. Yu Zhang was in charge of data analysis. Yi Zhang, Yawei Zhang, Xiaoying Ji, Chunxia Zhou, Lizhong Xie, Liming Cao, Niangui Zhao were responsible for collecting and analyze audio data. Jianfeng Cai, Yixiang Ye and Qifeng Wei are the member of Conv-MG. Shuisheng Wu, Wangyue Wang, Wen Xu, Yiqun Hu, Wensheng Pan, Yulong He, Changhua Zhang, Jian Chen are the consultant for Conv-MG. Jianping Jiang, Yongchao Hu, Chao Zheng, Hang Xiao were participated in equipment debugging and technical support. Jianquan He, Wen Zhang, Yunhong Lei, Zhenghua Jiang, Weiping Hu, Wenjuan Qin, Wanyu Wang, Xiaofang Zheng has given important help in starting the research. According to the author ranking, Wenjuan Qin and the previous authors are co-first authors.

## Declaration of interests

We declare no competing interests.

## Notes

### Competing Interest Statement

The authors have declared no competing interest.

